# Forensic Alcohol Calculations in transgender individuals undergoing gender-affirming hormonal treatment

**DOI:** 10.1101/2022.02.16.22271035

**Authors:** Peter D Maskell, Katherine J C Sang, Steven B Heymsfield, Sue Shapses, Alanna de Korompay

## Abstract

There are an increasing number of individuals undergoing gender-affirming hormonal treatment (GAHT) to treat gender dysphoria. Forensic alcohol calculations require knowledge of the sex of the individual but this may disadvantage trans people as research has demonstrated that there are physiological changes in individuals who are undergoing GAHT.

Using previously published studies on total body water (TBW) in cis individuals and the known changes in lean body mass and hematocrit in trans individuals, it is possible to revise the rubric for the estimation of TBW in trans individuals. We propose these revised equations are used for transgender individuals. When using these revised rubrics, we determined that for trans women the use of the cis male (sex at birth) anthropometric TBW equation only gives a small underestimation of TBW (0.9%) compared to the underestimation (−17.7%) of TBW using the cis female (affirmed gender) TBW equation. For trans men the use of gender at birth (cis female) TBW equation gives the largest disadvantage, underestimating TBW by -10.8 % compared to the affirmed gender (cis male) TBW equation that over estimates TBW by 6.6 %. For this reason, we recommend that if the sex at birth of an individual is not known or disclosed, any forensic alcohol calculations in the report are made assuming that the gender declared by the individual is their sex at birth.

**HIGHLIGHTS:**

- Large number of people taking gender-affirming hormonal treatment (GAHT) around the world
- Total body water is altered in individuals taking GAHT
- Forensic alcohol calculation results are affected by GAHT
- Recommended that in an individual undergoing GAHT revised Watson equation used
- Forensic alcohol reports state assumption that individual is cis (unless sex at birth is known)

---

Around the world there are estimated to be between 0.1 % and 2 % of the population who are transgender (1). Transgender is defined as people who have a gender identity which differs from the sex they were assigned at birth (1). The true number of transgender individuals around the world is unclear and is most likely being underestimated due to cultural sensitivities (2). Based on current data, around 80% of transgender individuals are either taking or want to take gender-affirming hormone therapy (GAHT) (3). The use of GAHT aims to align the characteristics of an individual with their gender identity. GAHT transgender women commonly receive oestrogen, often in conjunction with an androgen blocker or gonadotrophin-releasing hormone analogues. GAHT transgender men receive testosterone (4,5). These treatments are known to alter the body characteristics of the individuals taking them (4,5) and these body changes may influence the results of forensic alcohol calculations that are often based on the sex of an individual rather than their gender. In forensic science it is important to have a rigorous evidence base for forensic practices, particularly making sure that the practices do not disadvantage individuals or groups of individuals that may lead to miscarriages of justice (6). Forensic alcohol calculations, probably the most performed forensic calculations, have a solid evidence base due to many years of research (summarised in (7)). However, as far as the authors are aware there are no published guidelines, or recommendations, for forensic alcohol calculations that take into account the body changes that occur in individuals undergoing GAHT. The United Kingdom Association of Forensic Toxicologists (UKIAFT) alcohol calculation guidelines do state that the information collected for forensic alcohol calculations should include “sex at birth” (8). The assumption in these guidelines is that the individual undergoing GAHT will have a total body water similar to individuals of the sex they were assigned at birth. However, to date there are no studies looking at the body changes in transgender individuals with regard to forensic alcohol calculations. Additionally, depending on the legal jurisdiction, if an individual has legally changed their gender, they are under no obligation to disclose their sex at birth. The aim of this study is to determine the best approach with regards to forensic alcohol calculations in individuals that are undergoing GAHT.

## Forensic Alcohol Calculations

The most common form of the equation, known as the Widmark equation, to estimate the blood alcohol concentration of an individual after consumption of a known amount of alcohol is:

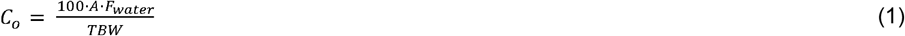

C_o_ – the hypothetical BAC at time zero before any metabolism has occurred (mg/100mL)

A – amount of pure ethanol consumed (g)

F_water_ – fraction of blood volume that is water (% w/v)

TBW – total body water of an individual (L)

For an individual undergoing GAHT there will be various physiological changes to their body. In the case of forensic alcohol calculations, the two variables that are likely to be altered by GAHT therapy are TBW and F_water_.

## Total body water (TBW) and gender-affirming hormonal treatment (GAHT)

There have been a number of studies looking at the changes of both body fat and fat free mass in individuals undergoing GAHT. These studies have shown that on average following the commencement of GAHT in trans women (assigned male at birth), there is an increase in body weight, an increase in body fat, and a decrease in lean body mass (9). On average in trans men (assigned female at birth), there is a decrease in body weight, decrease in body fat, and an increase in lean body mass following the commencement of GAHT (9). The variable of importance here for forensic alcohol calculations is that of lean body mass. Lean body mass is proportional to total body water, as the water content of the tissues is considered a constant (10,11). Thus, if the changes of lean body mass following GAHT are known the changes in total body water after GAHT can be estimated. The revised TBW in transgender individuals can then be utilised in forensic alcohol calculations. In a meta-analysis of individuals undergoing GAHT lean body mass was observed in trans women, on average, to decrease by -2.44 kg (−2.76 to -2.11 kg; 95 % CI) and on average, to increase in trans men by 3.87 kg (3.22 to 4.53 kg; 95 % CI) (9). In a study of 179 trans women and 162 trans men one year (12 months) after commencement of GAHT, lean body mass had decreased by -3 % [-4 to -2 %; 95 % CI] in trans women and increased by +10 % [9 to 11 %; 95 % CI] in trans men (12). Thus, the mean change in TBW for trans men would be approximately +10 % and approximately -3% in trans women (FIG. 1).

**Figure 1:**
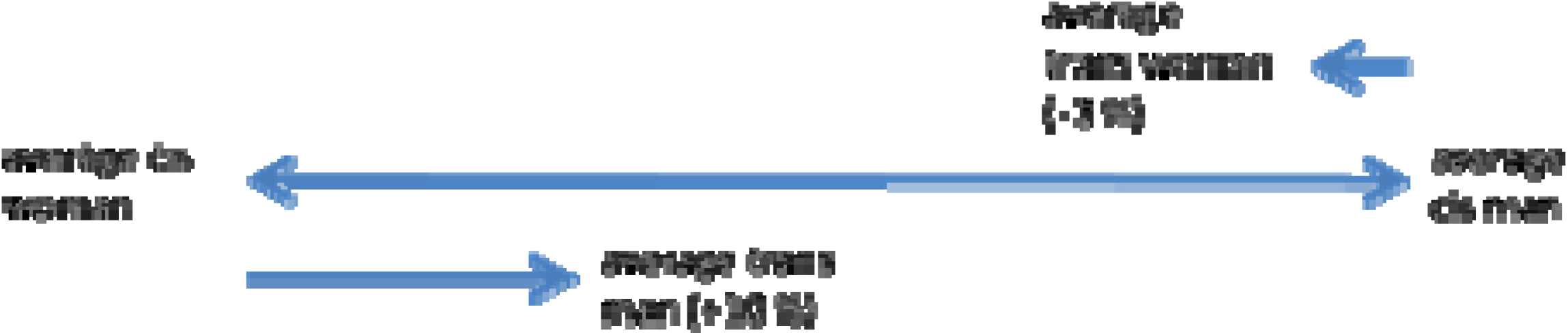
Mean percentage differences in total body water between cis men and cis women and the percentage change in total body water in trans people following gender affirming hormone therapy (GAHT).

## Percentage of blood that is water (F_water_) and gender-affirming hormonal treatment (GAHT)

Forensic alcohol calculations to determine the blood alcohol concentration of an individual after consuming a known amount of ethanol rely on not only the estimation of the individuals TBW, but also F_water_. Taking into account any potential changes in the percentage of blood that is water in people undergoing GAHT is also important. To date no studies have been directly carried out on the individuals undergoing GAHT and their F_water_, however previous work has demonstrated that whole blood water (WBW) correlates (r = -0.96) with hematocrit (Hct) (13,14) using the equation (2).

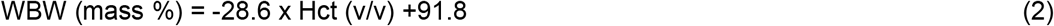

If the change in Hct was known in trans individuals the change in F_water_ could then be estimated. Studies in individuals undertaking GAHT have shown that there is on average a decrease in hematocrit in trans women and an increase in hematocrit in trans men (15). In a study of 239 trans women undergoing GAHT, hematocrit decreased from 45.1 Hct % [42.7– 47.59 Hct %; 95 % CI] at baseline to 41.0 Hct % [39.9-43 Hct %; 95 % CI]; a change of -4.1 Hct % (3.50-4.37 Hct %; 95 %CI) stabilising after 3 months. In the same study looking at 192 trans men undergoing GAHT the hematocrit increased by 4.9 Hct % [3.82-5.25 Hct %; 95 % CI] from 41.1 Hct % [39.0-42.6 Hct %; 95% CI] to 46.0 Hct % [44.0-47.0 Hct %; 95 % CI] after 12 months (15). After the conversion of hematocrit to WBW using equation (2), WBW (mass %) is 80.07 for trans women and 78.64 in trans men. WBW then needs to be converted to F_water_, (%w/v) by multiplying the blood water content percentage by 1.0506, the specific gravity of blood at 37 °C (16). This gives an F_water_ of 0.841 for trans women (17) compared to 0.838 for cis (gender identity equal to sex assigned at birth) women and 0.826 for trans men compared to 0.825 for cis men (17).

## Changes in total body water (TBW) in individuals undergoing gender affirming hormone therapy (GAHT)

The studies above have shown that there are body changes that will alter the results of forensic alcohol calculations in transgender individuals when compared to cisgender individuals. The work of Klaver et al (12) demonstrated that on average, the mean change in TBW for trans men was approximately +10 % and approximately -3% in trans women. As can be seen in Table 1 the actual change in lean body mass differs according to the body mass index (BMI) of the individual before the start of GAHT. In order to investigate the effects of these changes on the estimation of TBW and C_o_ in transgender individuals, two data sets were utilised: 1) the assigned male (cis male) at birth data (n = 582) and 2) the assigned female (cis female) at birth (n = 884) data. Both of these data sets are part of a study from the New York Obesity Research Centre at St. Lukes-Roosevelt Hospital, New York measure as part of a clinical study (18,19). These data sets comprised of the sex at birth, height, body mass (weight), age, and total body water (measured by the ^3^H-dilution method). Table 2 shows the mean (± SD) total body water ranges grouped into BMI of both cis men and cis women. Based on these data, cis males have a mean TBW of 45.9 ± 7.4 L (n = 582) and cis women have a mean TBW of 33.9 ± 6.2 L (n = 884) with a mean difference of 35.4 % (∼12 L). The measured TBW of the cis males and cis females was revised to estimate the TBW of trans individuals based on percentage changes in lean body mass based on the BMI of the cis individuals, detailed in Table 1. As can be seen in Table 3 the mean (± SD) TBW of trans women based on the starting cis male population is estimated to be 44.3 ± 7.0 L and for trans men to be 37.3 ± 6.4 L based on a starting cis female population. The only current guidance that applies to alcohol calculations in trans individuals suggests that the gender at birth should be used for ethanol calculations (8). However, based on the data from Klaver et al (12), on average the TBW of a transwoman would be 3 % lower than a cis man and 10 % higher for a cis woman compared to a trans man. Overall, as shown in table 3, there is a +30.7 % difference between the average TBW of trans woman compared to the average TBW of a cis woman with a -18.7% mean difference in the average TBW of a trans man compared to the average TBW of a cis man. Although there is a change in physiological parameters (TBW) towards that of the chosen rather than assigned sex at birth, the physiological changes after undergoing GAHT are not as large as would be expected if the individual has been assigned that gender at birth.

**Table 1:**
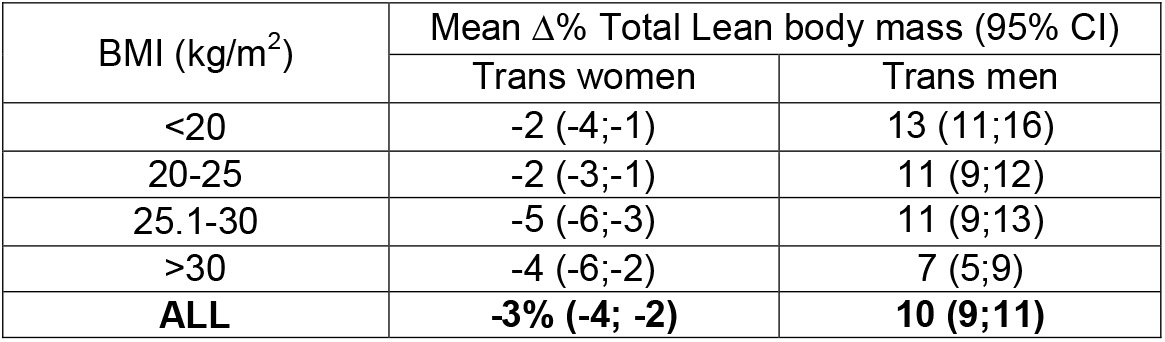
Changes in total lean body mass based on body mass index (BMI) of people undergoing gender-affirming hormonal treatment (GAHT) 1 year post the start of therapy. Data from Klaver et al. (12).

| BMI (kg/m<sup>2</sup>) | Mean Δ% Total Lean body mass (95% CI) |  |
| --- | --- | --- |
|  | Trans women | Trans men |
| <20 | -2 (-4;-1) | 13 (11;16) |
| 20-25 | -2 (-3;-1) | 11 (9;12) |
| 25.1-30 | -5 (-6;-3) | 11 (9;13) |
| >30 | -4 (-6;-2) | 7 (5;9) |
| **ALL** | **-3% (-4; -2)** | **10 (9;11)** |

**Table 2:** Mean total body water of cis men and cis women based on body mass index (BMI) based on data from Maskell et al (22).

| BMI (kg/m<sup>2</sup>) | Cis man (TBW (L)) |  |  | Cis woman (TBW (L)) |  |  | % Difference in mean TBW in cis men compared to cis women |
| --- | --- | --- | --- | --- | --- | --- | --- |
|  | mean | SD | n | mean | SD | n |  |
| <20 | 36.2 | 4.4 | 15 | 28.9 | 3.4 | 73 | 25.3 |
| 20-25 | 43.1 | 5.9 | 271 | 31.6 | 4.3 | 334 | 36.4 |
| 25.1-30 | 46.9 | 6.1 | 205 | 33.1 | 4.0 | 238 | 41.7 |
| >30 | 53.5 | 8.1 | 91 | 39.5 | 7.2 | 239 | 35.4 |
| ALL | 45.9 | 7.4 | 582 | 33.9 | 6.2 | 884 | 35.4 |

**Table 3:** Estimated mean Total Body Water (TBW) of transgender individuals after 12 months of gender-affirming hormonal treatment (GAHT) using data from Maskell et al(22) **assuming the GAHT caused changes TBW according to data collected by Klaver et al (12)**.

|  | Trans women (TBW (L)) |  |  | Trans men (TBW (L)) |  |  | % Difference in mean TBW between trans women and cis women | % Difference in mean TBW in trans men compared cis men | % Difference in mean TBW trans men compared to trans women |
| --- | --- | --- | --- | --- | --- | --- | --- | --- | --- |
| BMI | mean | SD | n | mean | SD | n |  |  |  |
| <20 | 35.5 | 4.3 | 15 | 32.7 | 3.9 | 73 | 22.8 | -9.7 | -7.9 |
| 20-25 | 42.2 | 5.7 | 271 | 35.1 | 4.7 | 334 | 33.5 | -18.6 | -16.8 |
| 25.1-30 | 44.6 | 5.8 | 205 | 36.7 | 4.5 | 238 | 34.7 | -21.7 | -17.7 |
| >30 | 51.3 | 7.8 | 91 | 42.3 | 7.7 | 239 | 29.9 | -20.9 | -17.5 |
| ALL | 44.3 | 7.0 | 582 | 37.3 | 6.4 | 884 | 30.7 | -18.7 | -15.8 |

## Revision of Anthropometric equations for individuals undergoing gender affirming hormone therapy (GAHT)

Based on the known changes to TBW in individuals undergoing GAHT, the anthropometric TBW equation of Watson et al (20) can be altered to take into account the effects of GAHT. The revised equations being:

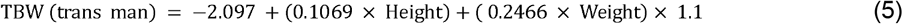

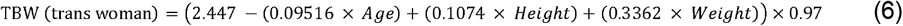

Weight (body mass in kg), Age (years), Height (cm).

The multiplications at the end of the trans men and trans women equations are adjusting for the mean change in TBW based on the studies above, +10 % in trans men and -3 % in trans women.

## Are trans individuals undergoing gender affirming hormone therapy (GAHT) disadvantaged by current practices?

### a) Total Body Water

The proposed best practice for the determination of TBW for transgender individuals undergoing GAHT would be to use the revised TBW equations (equation 5 and 6). Current UKIAFT guidelines for forensic practitioners state that the individuals total body water should be determined based on the individual’s sex at birth. Complications to this approach include that the transgender individual undergoing GAHT may have legally changed their gender and decide not to disclose their sex at birth for use in forensic alcohol calculations. This may bias the calculated result of the forensic alcohol calculation. Using the data from (18,19) we calculated the “true” mean total body water of trans men (equation 5) and trans women (equation 6). These data then allowed the calculation of the percentage difference in TBW if a) the cis male and b) the cis female TBW equations were used. As can be seen in Table 4 on average the TBW of a trans man would be underestimated by, on average, 4.2 L (10.8 % difference) if the female (sex at birth) TBW equation was used to estimate TBW. TBW would be overestimated, on average, by 2.4 L (6.6 % difference) if the male (affirmed gender) TBW equation was used. In transwomen the TBW would be underestimated, on average, by 0.6 L (0.9 % difference) if the male (sex at birth) TBW equation was used to estimate the individuals TBW. TBW would be under estimated, on average, by 8.1 L (17.7 % difference) if the female TBW (affirmed gender) equation was used. These data demonstrate that trans women would be most disadvantaged if the cis female (affirmed gender) TBW anthropometric equation was used to estimate their TBW. Trans men would be most disadvantaged if the cis female (sex at birth) anthropometric TBW equation was used to estimate their TBW.

**TABLE 4:** Difference in mean TBW for transgender individuals* when the TBW is estimated with the proposed trans gender equation compared to a) TBW being estimated using sex at birth TBW equation or b) TBW is estimated using the affirmed gender TBW equation.

| BMI | Trans women |  |  |  | Trans men |  |  |  |
| --- | --- | --- | --- | --- | --- | --- | --- | --- |
|  | Difference in TBW between sex at birth (cis man) and trans gender (trans woman) (L) | a) % TBW between sex at birth (cis man) and trans gender (trans woman) | b) Difference in TBW between affirmed gender (cis woman) and trans gender (trans woman) (L) | b) % Difference in TBW between affirmed gender (cis woman) and trans gender (trans woman) | a) Difference in TBW between sex at birth (cis woman) and trans gender (trans man) (L) | a) % Difference in TBW between sex at birth (cis woman) and trans gender (trans man) | b) Difference in TBW between affirmed gender (cis man) and trans gender (trans man) (L) | b) % Difference in TBW between affirmed gender (cis man) and trans gender (trans man) |
| <20 | -0.3 | -0.6 | -5.5 | -15.0 | -5.0 | -14.6 | 0.5 | 2.0 |
| 20-25 | -1.6 | -3.2 | -8.5 | -19.3 | -5.3 | -14.2 | 0.4 | 1.8 |
| 25.1-30 | -0.3 | 0.0 | -7.9 | -16.8 | -4.1 | -10.5 | 1.9 | 5.6 |
| >30 | 1.9 | 4.1 | -8.2 | -15.4 | -2.5 | -5.2 | 6.5 | 15.6 |
| ALL | -0.6 | -0.9 | -8.1 | -17.7 | -4.2 | -10.8 | 2.4 | 6.6 |

\* After 12 months of gender-affirming hormonal treatment (GAHT)

### Estimated BAC at time zero (C_o_)

It is important to determine how these differences in TBW observed in transgender individuals undergoing GAHT, described above, would alter the calculated C_o._ In order to look at the differences in C_o._ we used equation 1 to calculate the C_o_. This calculation used the TBW calculated above, the F_water_ of 0.838 (cis women); 0.825 (cis man); 0.841 (trans women) and 0.826 (trans men). Finally, we used 2 different doses of ethanol. It was assumed that an individual had consumed either 2 UK units of alcohol (16 g; ∼ 2 × 25 ml “shots” of 40% ABV vodka) or 10 UK units of ethanol 80 g (∼ 1 × 750 ml bottle of wine, 13% ABV). Table 5 shows the calculated C_o_, with table 6 showing the mean and percentage differences in C_o_ between the various groups. As with the TBW the estimation of C_o_ in transgender women using the cis female (assigned gender) TBW anthropometric equation would disadvantage trans women the most with a mean overestimation of C_o_ of ∼ 7 mg/100ml (∼ 23 % difference) for a dose of 16 g of ethanol and a mean overestimation of C_o_ of ∼ 32 mg/100ml (∼ 21 % difference) for a dose of 80 g of ethanol. For trans men, as with TBW the use of the cis female (sex at birth) TBW equation would give the greatest disadvantage with a mean overestimation of C_o_ of ∼ 6 mg/100ml (∼17 % difference) for a dose of 16 g of ethanol. For an 80 g dose of ethanol there would be a mean overestimation of C_o_ of ∼26 mg/100ml (∼14 % difference) for trans men if the cis female (sex at birth) calculations were used. Overall trans men would be disadvantaged to a greater extent with the use of sex at birth than trans women.

**Table 5:** Mean estimated Blood Alcohol concentration at time zero (C_o_) after the consumption of 16 g or 80 g of ethanol (alcohol) for transgender individuals* when a) the sex at birth; b) affirmed gender or c) specific transgender calculations are used.

|  |  | Mean Estimated C<sub>0</sub> (mg/100ml) |  |  |  |  |  |
| --- | --- | --- | --- | --- | --- | --- | --- |
|  |  | Trans women |  |  | Trans men |  |  |
| Dose of Ethanol (g) | BMI (kg/m<sup>2</sup>) | Cis Male calculation (sex at birth) | Trans female equation | Cis female calculation (affirmed gender) | cis female calculation (sex at birth) | trans male calculation | cis male calculation (affirmed gender) |
| **16** | <20 | 38 | 38 | 45 | 49 | 41 | 40 |
|  | 20-25 | 33 | 33 | 40 | 45 | 38 | 38 |
|  | 25.1-30 | 30 | 31 | 37 | 41 | 37 | 35 |
|  | >30 | 25 | 27 | 32 | 35 | 32 | 28 |
|  | ALL | 31 | 31 | 38 | 42 | 36 | 34 |
| **80** | <20 | 190 | 192 | 225 | 243 | 205 | 201 |
|  | 20-25 | 164 | 163 | 200 | 226 | 192 | 188 |
|  | 25.1-30 | 150 | 153 | 184 | 207 | 183 | 173 |
|  | >30 | 127 | 134 | 158 | 172 | 161 | 140 |
|  | ALL | 154 | 156 | 188 | 208 | 182 | 172 |

**\* After 12 months of gender-affirming hormonal treatment (GAHT)**

**TABLE 6:** Difference in mean C_o_ for transgender individuals* when the C_o_ is calculated with the proposed trans gender equation compared to a) C_o_ is calculated using sex at birth or b) C_o_ is calculated using the assigned gender.

|  |  | Estimated C<sub>0</sub> (mg/100ml) |  |  |  |  |  |  |  |
| --- | --- | --- | --- | --- | --- | --- | --- | --- | --- |
|  |  | Trans women |  |  |  | Trans men |  |  |  |
| Dose of Ethanol (g) | BMI | Difference in C<sub>0</sub> between sex at birth (cis man) and trans gender (trans woman) (mg/100ml) | a) % Difference in C<sub>0</sub> between sex at birth (cis man) and trans gender (trans woman) | b) Difference in C<sub>0</sub> between affirmed gender (cis woman) and trans gender (trans woman) (mg/100ml) | b) % Difference in C<sub>0</sub> between affirmed gender (cis woman) and trans gender (trans woman) | a) Difference in TBW between sex at birth (cis woman) and trans gender (trans man) (L) | a) % Difference in TBW between sex at birth (cis woman) and trans gender (trans man) | b) Difference in TBW between affirmed gender (cis man) and trans gender (trans man) (L) | b) % Difference in TBW between affirmed gender (cis man) and trans gender (trans man) |
| 16 | <20 | 0 | 0.0 | 7 | 18.4 | 8 | 19.5 | -1 | -2.4 |
|  | 20-25 | 0 | 0.0 | 7 | 21.2 | 7 | 18.4 | 0 | 0.0 |
|  | 25.1-30 | -1 | -3.2 | 6 | 19.4 | 4 | 10.8 | -2 | -5.4 |
|  | >30 | -2 | -6.3 | 5 | 18.5 | 3 | 9.4 | -4 | -12.5 |
|  | ALL | 0 | -0.6 | 7 | 22.6 | 6 | 16.7 | -2 | -5.6 |
| 80 | <20 | -2 | -1.0 | 33 | 17.2 | 38 | 18.5 | -4 | -2.0 |
|  | 20-25 | 1 | 0.8 | 37 | 22.7 | 34 | 17.7 | -4 | -2.1 |
|  | 25.1-30 | -3 | -1.8 | 31 | 20.3 | 24 | 13.1 | -10 | -5.5 |
|  | >30 | -7 | -5.4 | 24 | 17.9 | 11 | 6.8 | -21 | -13.0 |
|  | ALL | -2 | -1.3 | 32 | 20.5 | 26 | 14.3 | -10 | -5.5 |

**\*After 12 months of gender-affirming hormonal treatment (GAHT)**

## What are the implications of using the affirmed gender rather than sex at birth for individuals undergoing gender affirming hormone treatment

Transgender individuals undergoing GAHT who have legally changed their gender from that assigned at birth are commonly under no obligation to give their sex at birth. Ideally for trans individuals the best estimation of TBW would be to use the revised anthropometric TBW equations. However, for trans women the use of the cis male (sex at birth) anthropometric TBW equation only gives a small disadvantage (TBW -1 %; C_o_ ∼-1 %) compared to the cis female (affirmed gender) equation (TBW -17.7 %; C_o_ ∼22 %). For trans men the use of sex at birth gives the largest disadvantage, cis female TBW equation (TBW -10.8 %; C_o_ ∼15 %) compared to the affirmed gender (cis male) TBW equation (TBW ∼6.6 %; C_o_ ∼5.5 %). For this reason, we recommend that if the sex at birth of an individual is not known or not disclosed any forensic alcohol calculations in the report are made assuming that the gender declared by the individual is their sex at birth.

### Ethanol elimination rates

The rate of ethanol elimination is also an important parameter in forensic alcohol calculations (7). However as the same ethanol elimination rates and ranges are used for both sexes in forensic ethanol calculations trans specific ethanol elimination rates and ranges do not need to be used (21).

### Limitations

This study is based on the mean changes that occur in TBW and F_water_ following 12 months of GAHT in trans men and trans women that are mainly Western Caucasians with an age range of 18 to 66. In order to validate TBW water equations for use in transgender individuals’ studies need to be carried out in transgender individuals of a wide range of ages, BMI, races and gender to develop anthropometric equations. Studies should also be carried out to determine the F_water_ in transgender individuals. Until these studies are carried out the equations detailed above give the best available forensic alcohol calculations for use in trans gender people who have completed a year of cross-sex hormonal therapy.

## Conclusions

This study has demonstrated that alcohol calculations for trans gender people undergoing GAHT should be carried out using the revised Watson et al. equations taking into account the changes in TBW and F_water_ that occur after cross-sex hormonal therapy, when the individual has declared they are trans gender. If it is not known if the individual is cis gender or trans gender then the forensic alcohol calculation report should state the assumption that the gender given by the individual is considered to be the sex at birth of the individual. The use of these revised equations and assumptions mean that transgender people are less likely to be disadvantaged in cases where forensic alcohol calculations need to be carried out.

## Data Availability

All data produced in the present study are available upon reasonable request to the authors

## Acknowledgements

The authors would like to thank Fiona Flaherty for her advice on rubrics and Jennifer Limoges for her constructive comments on the draft manuscript.

## References

1. Deutsch MB. Making It Count: Improving Estimates of the Size of Transgender and Gender Nonconforming Populations. LGBT Heal 2016;3(3):181–5. https://doi.org/10.1089/lgbt.2016.0013.

2. Goodman M, Adams N, Corneil T, Kreukels B, Motmans J, Coleman E. Size and Distribution of Transgender and Gender Nonconforming Populations: A Narrative Review. Endocrinol Metab Clin North Am 2019;48(2):303–21. https://doi.org/10.1016/j.ecl.2019.01.001.

3. Grant JM, Mottet LA, Tanis J, Harrison J, Herman JL, Keisling M. National transgender discrimination survey report on health and health care. 2010. <https://cancer-network.org/wp-content/uploads/2017/02/National_Transgender_Discrimination_Survey_Report_on_health_and_health_care.pdf (accessed november 21,2021).

4. Irwig MS. Testosterone therapy for transgender men. lancet Diabetes Endocrinol 2017;5(4):301–11. https://doi.org/10.1016/S2213-8587(16)00036-X.

5. Tangpricha V, den Heijer M. Oestrogen and anti-androgen therapy for transgender women. lancet Diabetes Endocrinol 2017;5(4):291–300. https://doi.org/10.1016/S2213-8587(16)30319-9.

6. President’s Council of Advisors on Science and Technology (PCAST). Forensic Science in Criminal Courts: Ensuring Scientific Validity of Feature-Comparison Methods. 2016. https://obamawhitehouse.archives.gov/sites/default/files/microsites/ostp/PCAST/pcast_forensic_science_report_final.pdf (accessed november 21, 2021).

7. Jones AW. Pharmacokinetics of Ethanol. In: Jones AW, Mørland JG, Liu RH, editors. Alcohol, Drugs, and Impaired Driving. CRC Press, 2020;275–346.

8. UKIAFT Guidelines for Alcohol Calculations Version 4.3. 2021. http://www.ukiaft.co.uk/image/catalog/documents/ukiaft-atd-v4.3.pdf (accessed november 21, 2021).

9. Klaver M, Dekker MJHJ, de Mutsert R, Twisk JWR, den Heijer M. Cross-sex hormone therapy in transgender persons affects total body weight, body fat and lean body mass: a meta-analysis. Andrologia 2017;49(5). https://doi.org/10.1111/and.12660.

10. Wang Z, Deurenberg P, Wang W, Pietrobelli A, Baumgartner RN, Heymsfield SB. Hydration of fat-free body mass: new physiological modeling approach. Am J Physiol 1999;276(6):E995–1003. https://doi.org/10.1152/ajpendo.1999.276.6.E995.

11. Wang Z, Deurenberg P, Wang W, Pietrobelli A, Baumgartner RN, Heymsfield SB. Hydration of fat-free body mass: review and critique of a classic body-composition constant. Am J Clin Nutr 1999;69(5):833–41. https://doi.org/10.1093/ajcn/69.5.833.

12. Klaver M, de Blok CJM, Wiepjes CM, Nota NM, Dekker MJHJ, de Mutsert R, et al. Changes in regional body fat, lean body mass and body shape in trans persons using cross-sex hormonal therapy: results from a multicenter prospective study. Eur J Endocrinol 2018;178(2):163–71. https://doi.org/10.1530/EJE-17-0496.

13. Beijering RJ, Gips CH, Huizenga JR, Jager J, Mackor AJ, Salomons H, et al. Whole blood and plasma water in health and disease: longitudinal and transverse observations and correlations with several different hematological and clinicochemical parameters. Clin Chim Acta 1997;258(1):59–68. https://doi.org/10.1016/s0009-8981(96)06428-5.

14. Lijnema TH, Huizenga JR, Jager J, Mackor AJ, Gips CH. Gravimetric determination of the water concentration in whole blood, plasma and erythrocytes and correlations with hematological and clinicochemical parameters. Clin Chim Acta 1993;214(2):129–38. https://doi.org/10.1016/0009-8981(93)90105-d.

15. Defreyne J, Vantomme B, Van Caenegem E, Wierckx K, De Blok CJM, Klaver M, et al. Prospective evaluation of hematocrit in gender-affirming hormone treatment: results from European Network for the Investigation of Gender Incongruence. Andrology 2018;6(3):446–54. https://doi.org/10.1111/andr.12485.

16. Trudnowski RJ, Rico RC. Specific gravity of blood and plasma at 4 and 37 °C. Clin Chem 1974;20(5):615–6. https://doi.org/10.1093/clinchem/20.5.615.

17. Maskell PD, Cooper GAA. Uncertainty in Widmark equation calculations: Volume of distribution and body mass. J Forensic Sci 2020;65(5):1676–84. https://doi.org/10.1111/1556-4029.14447.

18. Levitt DG, Heymsfield SB, Pierson RN, Shapses SA, Kral JG. Physiological models of body composition and human obesity. Nutr Metab 2007;4(1):19. https://doi.org/10.1186/1743-7075-4-19.

19. Levitt DG, Heymsfield SB, Pierson RN, Shapses SA, Kral JG. Physiological models of body composition and human obesity. Nutr Metab 2009;6:7. https://doi.org/10.1186/1743-7075-6-7.

20. Watson PE, Watson ID, Batt RD. Total body water volumes for adult males and females estimated from simple anthropometric measurements. Am J Clin Nutr 1980;33(1):27–39. https://doi.org/10.1093/ajcn/33.1.27.

21. Jones AW. Evidence-based survey of the elimination rates of ethanol from blood with applications in forensic casework. Forensic Sci Int 2010;200(1–3):1–20. https://doi.org/10.1016/j.forsciint.2010.02.021.

22. Maskell PD, Jones AW, Heymsfield SB, Shapses S, Johnston A. Total body water is the preferred method to use in forensic blood-alcohol calculations rather than ethanol’s volume of distribution. Forensic Sci Int 2020;316:110532. https://doi.org/10.1016/j.forsciint.2020.110532.

